# Cerebral venous thrombosis and portal vein thrombosis: a retrospective cohort study of 537,913 COVID-19 cases

**DOI:** 10.1101/2021.04.27.21256153

**Authors:** Maxime Taquet, Masud Husain, John R Geddes, Sierra Luciano, Paul J Harrison

## Abstract

**Objectives:** To estimate the absolute risk of cerebral venous thrombosis (CVT) and portal vein thrombosis (PVT) in the two weeks following a diagnosis of COVID-19, and to assess the relative risks (RR) compared to influenza or the administration of an mRNA vaccine against COVID-19.

**Design:** Retrospective cohort study based on an electronic health records network

**Setting:** Linked records between primary and secondary care centres within 59 healthcare organisations, primarily in the USA

**Participants:** All patients with a confirmed diagnosis of COVID-19 between January 20, 2020 and March 25, 2021 were included (N=537,913, mean [SD] age: 46.2 [21.4] years; 54.9% females). Cohorts (matched for age, sex, and race) of participants diagnosed with influenza (N=392,424) or receiving the BNT162b2 or mRNA-1273 vaccine (N=366,869) were used for comparison.

**Main outcome measures:** Diagnosis of CVT (ICD-10 code I67.6) or PVT (ICD-10 code I81) within 2 weeks after a diagnosis of COVID-19.

**Results:** The incidence of CVT after COVID-19 diagnosis was 42.8 per million people (95% CI 28.5–64.2) including 35.3 per million (95% CI 22.6–55.2) first diagnoses. This was significantly higher than the CVT incidence in a matched cohort of patients with influenza (RR=3.83, 95% CI 1.56–9.41, P<0.001) and people who received an mRNA vaccine (RR=6.67, 95% CI 1.98–22.43, P<0.001). The incidence of PVT after COVID-19 diagnosis was 392.3 per million people (95% CI 342.8–448.9) including 175.0 per million (95% CI 143.0–214.1) first diagnoses. This was significantly higher than the PVT incidence in a matched cohort of patients with influenza (RR=1.39, 95% CI 1.06–1.83, P=0.02) and people who received an mRNA vaccine (RR=7.40, 95% CI 4.87–11.24, P<0.001). Mortality after CVT and PVT was 17.4% and 19.9% respectively.

**Conclusions:** The incidence of CVT and PVT is significantly increased after COVID-19. The data highlight the risk of serious thrombotic events in COVID-19 and can help contextualize the risks and benefits of vaccination in this regard.

**What is known:**

‐ A systematic review of cohort studies suggested an incidence of CVT among *hospitalised* patients with COVID-19 to be about 800 per million patients. There was evidence of selection, ascertainment, and reporting bias in all included studies.
‐ The incidence of CVT and PVT among both hospitalised and non-hospitalised patients with COVID-19 is unknown.
‐ It is unknown if COVID-19 increases the risk of CVT and PVT.

**What this study adds:** Our study estimates that the absolute risk of CVT and PVT are respectively 42.8 and 392.3 per million patients (both hospitalised and non-hospitalised) in the 2 weeks after a diagnosis of COVID-19. COVID-19 increases the risk of CVT and PVT compared to patients diagnosed with influenza, and to people who have received a COVID-19 mRNA vaccine.

---

There are concerns about a possible association between vaccines against SARS-CoV-2 and cerebral venous thrombosis (CVT, also called cerebral venous sinus thrombosis [1]). The concern has focused primarily on ChAdOx1 nCoV-19 (“Oxford-AstraZeneca”) vaccine, and more recently the Ad26.COV2.S (“Janssen”) vaccine. Emerging data suggest that the association reflects a ‘vaccine-induced thrombotic thrombocytopaenia’ (VITT) [2,3]. Governments and regulatory authorities have reacted by restricting the use of the two vaccines in different subgroups of the population, based on a risk-benefit analysis. Yet one key component of the risk-benefit calculation is currently unknown: the absolute risk of CVT following a diagnosis of COVID-19. To date there are only a few case series of CVT post-COVID-19, and a few cohort studies limited to hospitalised patients and with evidence of selection, ascertainment, and reporting bias [4].

Here, using an electronic health records network primarily based in the USA, we estimated the incidence of CVT occurring in confirmed COVID-19 cases (both hospitalised and non-hospitalised) and compared this incidence to two other groups: people who received a COVID-19 mRNA vaccine, and a cohort of patients with influenza. We could not make a direct comparison with rates after the ChAdOx1 nCoV-19 (“Oxford-AstraZeneca”) vaccine because this has not been used in the USA. We also examined the rate of portal vein thrombosis (PVT), another diagnosis associated with thrombosis in the venous system and thought to occur in VITT [3].

## Methods

### Data

We used TriNetX Analytics, a federated electronic health records network recording anonymized data from 59 healthcare organizations, primarily in the USA, totalling 81 million patients. For details, see [5], and the supplement.

The process by which the data is de-identified is attested to through a formal determination by a qualified expert as defined in Section §164.514(b)(1) of the HIPAA Privacy Rule, so that ethical approval from an institutional review board is not needed.

### Primary analysis

A cohort of all patients who had a confirmed diagnosis of COVID-19 (ICD-10 code U07.1) between January 20, 2020 and March 25, 2021 was defined for study. The absolute risk of a diagnosis of CVT (ICD-10 code I67.6) was calculated by identifying those patients in the cohort who had the diagnosis in the two weeks following their diagnosis of COVID-19. The absolute risk of PVT (ICD-10 code I81) was also calculated. For the whole COVID-19 cohort, and for cases with CVT or PVT following COVID-19, baseline characteristics are reported. We identified patients who had a reported high D-dimer (> 5mg/L), low fibrinogen (< 200 mg/dL), or thrombocytopenia within the 2 weeks after their COVID-19 diagnosis. We also assessed how many of them had died (and, if so, when) by the time of the analysis (April 21, 2021).

A causal link between COVID-19 and CVT/PVT cannot be established with a simple cohort study. However, a testable corollary of such a causal association is that the rate of new CVT/PVT diagnoses decreases with time from the index event. We tested this corollary by comparing the absolute risk within two weeks of diagnosis (Week 1 and 2) with the absolute risk with the next two weeks (Week 3 and 4) and the two weeks thereafter (Week 5 and 6). For this part of the analysis, only patients diagnosed on or before February 28, 2021 were included to allow for sufficient follow-up.

Two control cohorts based on other index events were used for comparison: a diagnosis of influenza (ICD-10 codes J09-J11) between January 20, 2018 and March 25, 2021 (an earlier start date was used for this event to achieve a sufficiently large sample), and receipt of a first dose of the two vaccines administered to this predominantly US population: the BNT162b2 (‘Pfizer-BioNTech’) vaccine or the mRNA-1273 (‘Moderna’) vaccine before March 25, 2021. We excluded from these cohorts any patients who had a diagnosis of COVID-19 on or after January 20, 2020.

These two cohorts were then matched to the cohort of patients with COVID-19 for age (defined as a continuous variable), sex (defined as a categorical variable taking the value: female, male, or other), and race (defined as a categorical variable taking the value: White, Black or African American, Asian, American Indian or Alaska Native, Native Hawaiian or Other Pacific Islander, or Unknown) using propensity score matching (see details below). Using these matched cohorts, we calculated the relative risk (RR) of a CVT diagnosis, a PVT diagnosis, and a diagnosis of thrombocytopenia in the two weeks after COVID-19 diagnosis compared to the other index events (i.e. influenza or vaccination).

### Secondary analyses

The analyses were repeated after broadening the diagnostic criteria for CVT to include I63.6 (cerebral infarction due to central thrombosis, non-pyogenic), G08 (intracranial and intraspinal phlebitis and thrombophlebitis), O22.5 (CVT in pregnancy) and O87.3 (CVT in the puerperium), in line with recent epidemiological studies that have taken this definition of CVT in other settings [6,7].

The analyses were also repeated after excluding those patients who had had a prior diagnosis of the event of interest (CVT or PVT).

### Statistical analyses

Fisher’s exact tests were used to compare characteristics (baseline and laboratory) and death rates between patients with COVID-19 who had a CVT (or PVT) compared to patients with COVID-19 who did not. Fisher’s exact tests were also used to test the null hypothesis that the relative risks of CVT and PVT in the two weeks after COVID-19 vs. influenza and vs. mRNA vaccine were equal to 1. Confidence intervals for absolute risks were based on Wilson score intervals. Confidence intervals for relative risks were based on Wald confidence limits, with Agresti-Coull adjustment to improve coverage when the risks are small [8].

Propensity score matching was used to create cohorts with matched baseline characteristics, and carried out within the TriNetX network. Propensity score 1:1 matching used a greedy nearest neighbour matching approach with a caliper distance of 0.1 pooled standard deviations of the logit of the propensity score. Any characteristic with a standardized mean difference (SMD) between cohorts lower than 0.1 is considered well matched. [9]

Statistical significance was set at a 2-sided P value < 0.05. Analyses were performed using R version 3.6.3. This study follows the STROBE reporting guidelines (see Supplement for a checklist). Further details about TriNetX, cohort definitions, and statistical analyses can be found in the supplement.

### Patient and Public Involvement

Patients and public were not involved in this study.

## Results

537,913 patients with a confirmed diagnosis of COVID-19 were included in this study (54.9% females, mean [SD] age 46.2 [21.4]; Table 1). Of these, 23 were diagnosed with a CVT in the two weeks following their diagnosis (absolute risk: 42.8 per million people, 95% CI 28.5–64.2, equivalent to an incidence of 111.5 per 100k person-years). The risk was significantly higher among patients with a history of cardiovascular diseases (Table 1), specifically arterial diseases, cerebral/precerebral artery stenosis/occlusion, and intracranial haemorrhage.

**Table 1.** Baseline characteristics of the whole COVID-19 cohort and the groups who received a diagnosis of CVT or PVT in the two weeks after COVID-19 diagnosis. The P-values from Fisher exact test (or t-test for age) for CVT and PVT groups compared to the rest of the COVID-19 cohort are shown.

|  | All patients<br>with COVID-19 | Patients with<br>COVID-19 and CVT |  | Patients with<br>COVID-19 and PVT |  |
| --- | --- | --- | --- | --- | --- |
|  | n (%) \| mean (SD) | n (%) \| mean (SD) | P | n (%) \| mean (SD) | P |
| **Sample size, n** | 537913 (100.0) | 23 (100.0) | - | 211 (100.0) | - |
| **Age, mean (SD), y** | 46.2 (21.4) | 46.5 (21.5) | 0.95 | 57.2 (14.6) | <0.001 |
| **Sex, n (%)** |  |  |  |  |  |
| Female | 295220 (54.9) | 16 (69.6) | 0.21 | 94 (44.5) | 0.0029 |
| Male | 242439 (45.1) | 7 (30.4) | 0.21 | 117 (55.5) | 0.0028 |
| **Race, n (%)** |  |  |  |  |  |
| White | 326258 (60.7) | 18 (78.3) | 0.091 | 145 (68.7) | 0.017 |
| Black | 93712 (17.4) | 4 (17.4) | 1 | 34 (16.1) | 0.72 |
| Asian | 14498 (2.7) | 2 (8.7) | 0.13 | 3 (1.4) | 0.39 |
| Other | 3909 (0.7) | 0 (0.0) | 1 | 1 (0.5) | 1 |
| Unknown | 99536 (18.5) | 2 (8.7) | 0.29 | 28 (13.3) | 0.051 |
| **Comorbidities at baseline, n (%)** |  |  |  |  |  |
| Obesity | 92903 (17.3) | 4 (17.4) | 1 | 57 (27.0) | <0.001 |
| Hypertension | 153305 (28.5) | 8 (34.8) | 0.49 | 115 (54.5) | <0.001 |
| CKD | 35582 (6.6) | 2 (8.7) | 0.66 | 51 (24.2) | <0.001 |
| Ischemic heart diseases | 48767 (9.1) | 5 (21.7) | 0.052 | 42 (19.9) | <0.001 |
| Cardiac failure | 29536 (5.5) | 2 (8.7) | 0.36 | 26 (12.3) | <0.001 |
| Arterial diseases | 38606 (7.2) | 7 (30.4) | <0.001 | 44 (20.9) | <0.001 |
| Venous diseases | 33222 (6.2) | 4 (17.4) | 0.05 | 171 (81.0) | <0.001 |
| Cerebral/Pre-cerebral artery stenosis | 20817 (3.9) | 6 (26.1) | <0.001 | 21 (10.0) | <0.001 |
| Intracranial haemorrhage | 4056 (0.8) | 5 (21.7) | <0.001 | 9 (4.3) | <0.001 |
| Dementia | 11872 (2.2) | 1 (4.3) | 0.4 | 3 (1.4) | 0.64 |
| Chronic lower resp. disease | 90617 (16.8) | 6 (26.1) | 0.26 | 69 (32.7) | <0.001 |
| Connective tissue disorders | 9671 (1.8) | 1 (4.3) | 0.34 | 15 (7.1) | <0.001 |
| Liver disease | 32972 (6.1) | 2 (8.7) | 0.65 | 148 (70.1) | <0.001 |
| Diabetes mellitus | 77038 (14.3) | 6 (26.1) | 0.13 | 73 (34.6) | <0.001 |
| Malignancy | 40636 (7.6) | 3 (13.0) | 0.25 | 100 (47.4) | <0.001 |
| Past CVT | 90 (0.02) | 4 (17.4) | <0.001 | 0 (0.0) | 1 |
| Past PVT | 676 (0.1) | 0 (0.0) | 1 | 117 (55.5) | <0.001 |

Among the 23 events, 7 were observed in patients under the age of 30, 4 between 30 and 39, 2 between 40 and 49, 3 between 50 and 59, 2 between 60 and 69, and 5 between 70 and 79. Four patients had also had a CVT diagnosed prior to their COVID-19 diagnosis, one between 4 and 8 weeks beforehand, and the other 3 more than 8 weeks prior. The incidence of CVT following COVID-19 significantly decreased with time from the index event (RR in 3^rd^ and 4^th^ week vs. first 2 weeks 0.24, 95% CI 0.098–0.59, P<0.001; RR in 5^th^ and 6^th^ week vs. first 2 weeks 0.12, 95% CI 0.036–0.40, P<0.001; Fig. 1).

**Figure 1.**
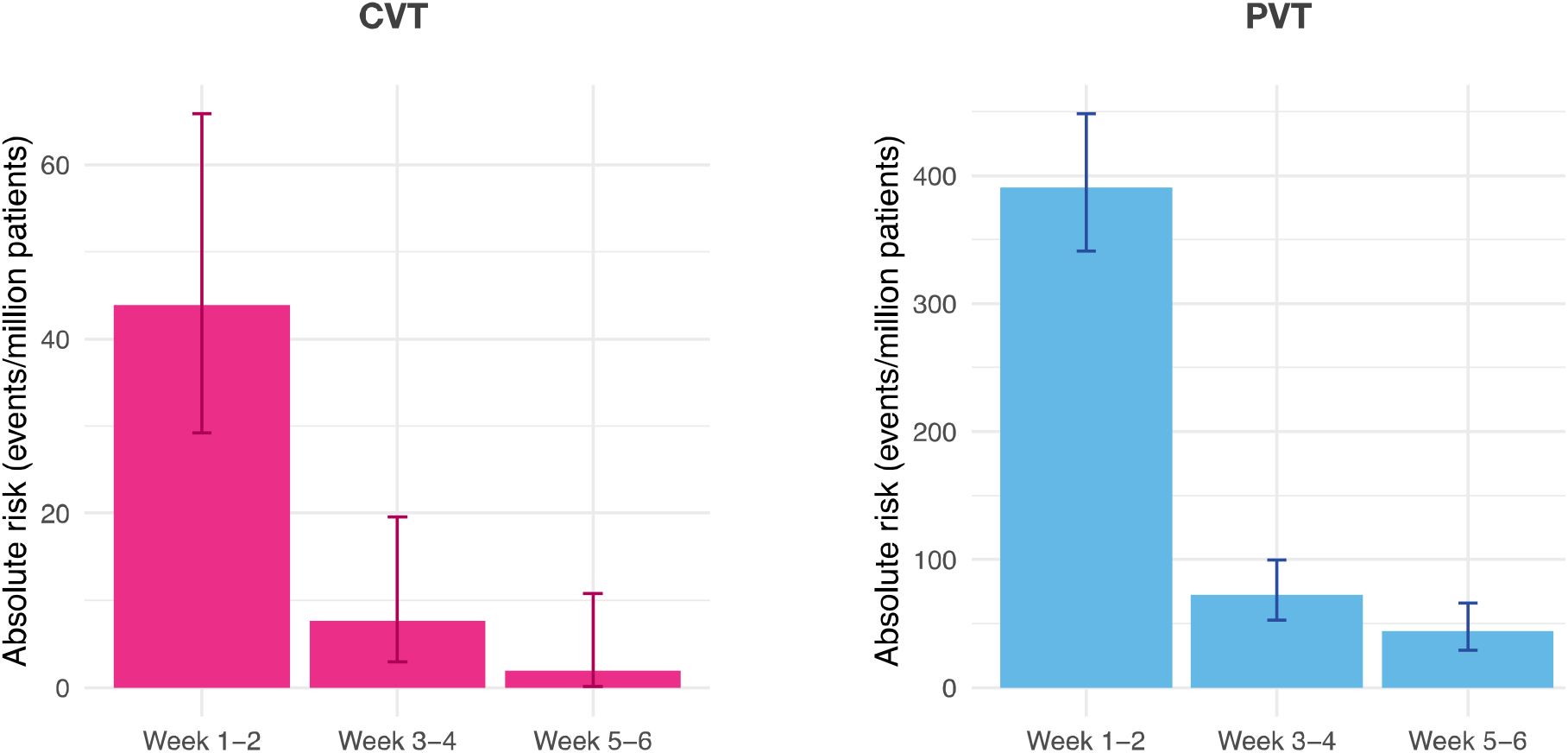
Incidence of CVT and PVT per million patients as a function of the time since diagnosis of COVID-19, from the first 2 weeks post diagnosis to the 5^th^ and 6^th^ week post diagnosis.

The absolute risk of PVT in the two weeks following COVID was 392.3 per million people (95% CI 342.8–448.9), equivalent to 1022.7 per 100k person-years. Among the 211 affected patients, 117 had a PVT prior to their COVID-19 diagnosis. The incidence of PVT following COVID-19 significantly decreased with time from the index event (RR in 3^rd^ and 4^th^ week vs. first 2 weeks 0.19, 95% CI 0.14–0.27, P<0.001; RR in 5^th^ and 6^th^ week vs. first 2 weeks 0.12, 95% CI 0.080–0.18, P<0.001; Fig. 1).

Laboratory data were available for a subset of the COVID-19 patients (Table 2). Although the data do not cover most patients with a diagnosis of CVT, they suggest that patients with CVT after COVID-19 were significantly more likely to have elevated D-dimer level than patients with COVID-19 who did not have CVT, whereas patients with PVT after COVID-19 were significantly more likely to have low fibrinogen level and thrombocytopenia. The death rate among patients with CVT in the two weeks after COVID-19 was 17.4% (4 out of 23 patients, 95% CI 6.98–37.1%; Figure S1 in the supplement) and that among patients with PVT after COVID-19 was 19.9% (42 out of 211 patients, 95% CI 15.1–25.8%; Figure S1 in the supplement) and were significantly higher than among patients with COVID-19 who did not have those events (P=0.005 for CVT and P<0.001 for PVT).

**Table 2.** Laboratory characteristics of the patients in each group. P values are from Fisher’s exact test, comparing the CVT and PVT groups to the rest of the COVID-19 cohort.

|  | All patients<br>with COVID-19 | Patients with<br>COVID-19 and CVT |  | Patients with<br>COVID-19 and PVT |  |
| --- | --- | --- | --- | --- | --- |
|  | n (%) | n (%) | P | n (%) | P |
| **D-dimer > 5 mg/L**<br>n/n with measurement (%) | 2047/69313 (3.0) | 2/6 (33.3) | 0.012 | 4/66 (6.1) | 0.14 |
| **Fibrinogen < 200 mg/dL**<br>n/n with measurement (%) | 1167/19602 (6.0) | 1/6 (16.7) | 0.31 | 22/46 (47.8) | <0.001 |
| **Thrombocytopenia**<br>(ICD-10 codes D69.49, D69.59,<br>D69.6) | 8840 (1.8) | 0 (0.0) | 1 | 59 (28.0) | <0.001 |
| **Death** | 16619 (3.1) | 4 (17.4) | 0.005 | 42 (19.9) | <0.001 |

The two-week risk of being diagnosed with a CVT was significantly higher in the cohort diagnosed with COVID-19 compared to a matched cohort diagnosed with influenza (N=392,424 in each cohort; RR=3.83, 95% CI 1.56–9.41, P<0.001; Fig. 2 and Table S1) or compared to a matched cohort receiving an mRNA vaccine (N=366,869 in each cohort; RR=6.67, 95% CI 1.98–22.43, P<0.001; Fig. 2 and Table S2). Similarly, the two-week risk of being diagnosed with a PVT was significantly higher in the cohort diagnosed with COVID-19 compared to a matched cohort diagnosed with influenza (RR=1.39, 95% CI 1.06–1.83, P=0.02; Fig. 2) or compared to a matched cohort receiving an mRNA vaccine (RR=7.40, 95% CI 4.87–11.24, P<0.001; Fig. 2). A diagnosis of thrombocytopenia was also significantly more likely in the two weeks after a diagnosis of COVID-19 than after a diagnosis of influenza (RR=1.23, 95% CI 1.18–1.28, P<0.001) or after receiving an mRNA vaccine (RR=30.8, 95% CI 27.1–35.0, P<0.001).

**Figure 2.**
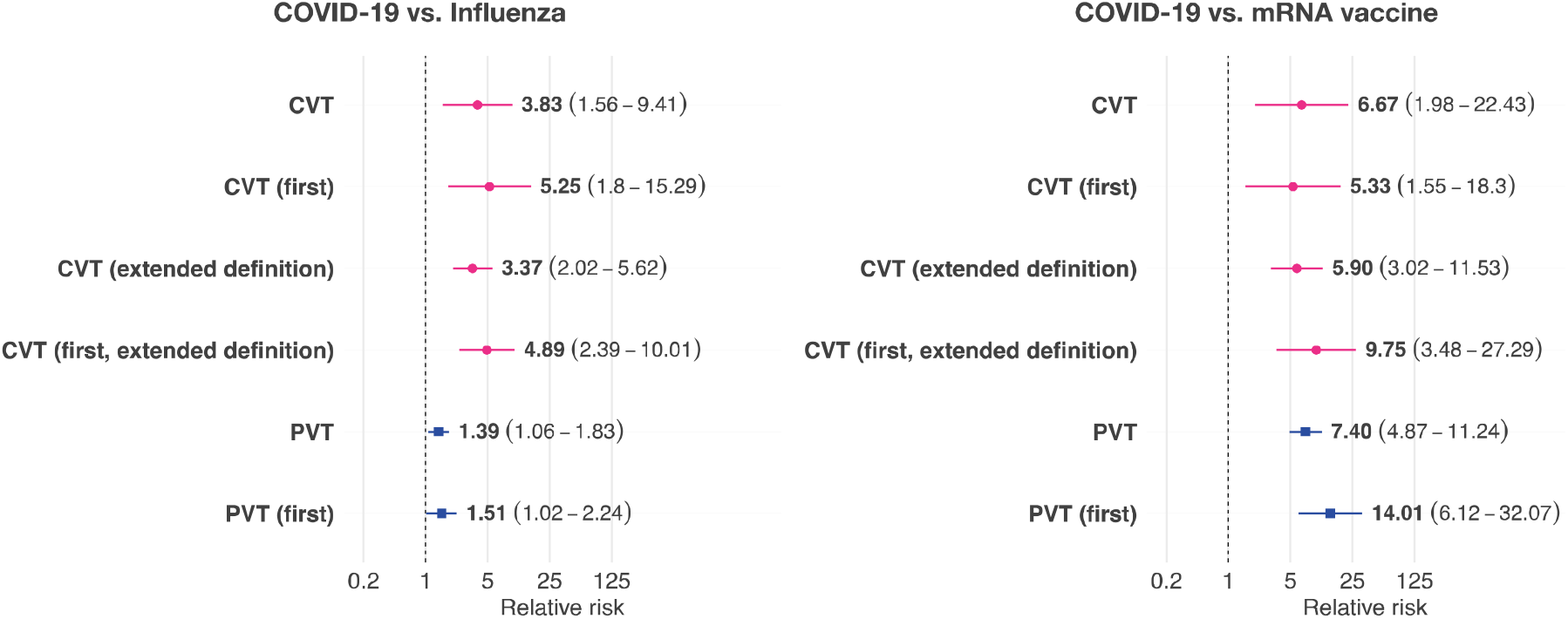
Relative risk of CVT and PVT after a diagnosis of COVID-19 compared to matched cohorts of people with a diagnosis of influenza (left) or receiving an mRNA vaccine (right). Horizontal lines and numbers in brackets represent the 95% confidence intervals.

When the definition of CVT in terms of ICD-10 codes was broadened, the incidence of CVT in the two weeks after COVID-19 was 148.7 per million people (95% CI 119.5–185.1), which was significantly higher than in the matched cohort of patients with influenza (RR=3.37, 95% CI 2.02–5.62; P<0.001) and the matched cohort of people receiving an mRNA vaccine (RR=5.90, 95% CI 3.02–11.53, P<0.001; Fig. 2). The majority of the extra cases came from the G08 diagnostic category.

Finally, we excluded patients who had also had a CVT or PVT prior to COVID-19. The incidence of CVT and PVT post-COVID-19 diagnosis were reduced accordingly (CVT: absolute risk 35.3 per million, 95% CI 22.6–55.2); PVT (absolute risk 175.0 per million, 95% CI 143.0–214.1), but all RRs were similar to those in the primary analysis (Fig. 2).

## Discussion

In a large electronic health records network, the absolute incidence of CVT and PVT in the 14 days after COVID-19 diagnosis was 42.8 and 392.3 per million patients respectively. The incidence rapidly decreased in the following weeks, which is compatible with a causal link between COVID-19 and those thrombotic events. However, causation cannot be demonstrated with the current study and residual confounding (e.g. increased medical monitoring directly after COVID-19 vs. a few weeks later) might contribute to this observation.

The incidence of CVT and PVT after COVID-19 is substantially greater than in the matched control cohorts. The incidence of CVT after a diagnosis of COVID-19 is also substantially greater than the expected incidence in the general population in the USA, estimated to be between 0.53 and 0.77 per million people in any 2-week period [7] and the rate is significantly higher than the highest of these estimates (binomial test: P<0.001).

The incidence is also many-fold higher than the latest reported incidence of CVT following administration of the first dose of the ChAdOx1 nCoV-19 (‘Oxford-AstraZeneca’) vaccine (reported by the European Medicines Agency to be around 5 per million vaccinated people [10]) and the latest reported incidence of CVT following administration of the Ad26.COV2.S (“J&J”) vaccine (reported by the Food and Drug Administration to be about 0.9 per million vaccinated people [11]).

The increased rate of CVT in COVID-19 is notable, being much more marked than the increased risks for other forms of stroke and cerebral haemorrhage [5]. The PVT data highlight that COVID-19 is associated with thrombotic events that are not limited to the cerebral vasculature.

Importantly, the present study cannot be used to draw conclusions on the relative risk of developing a CVT or PVT after receiving an mRNA vaccine compared to the baseline incidence or compared to other vaccines. Far larger samples are needed (such as those used by the EMA and the FDA pharmacovigilance studies) because the events have so far been found to be extremely rare. The observed incidence of CVT in the matched cohort of people who received an mRNA vaccine is compatible with even the lowest estimate of the baseline rate in the USA of 0.53 per million people in any 2-week period (binomial test: P=0.18 [7]).

The main strengths of this study are its large sample size and the use of matched cohorts. However, the study also has several limitations and results should be interpreted with caution. First, while cohorts were matched for age, sex, and race, they were not matched for comorbidities so that the latter might be contributing to the association between COVID-19 and subsequent CVT/PVT. Second, we have no information about diagnostic accuracy or completeness of records, though this is likely to be less of an issue for CVT or PVT compared to many diagnoses. Third, some cases of COVID-19, especially those early in the pandemic, are undiagnosed, and we cannot generalise our risk estimates to this population. Similarly, COVID-19 vaccines are also being delivered by sites which are not part of an HCO, and many of these may not be coded in the electronic health record, and so the relative risk estimates may not apply to them. Fourth, the absence of key haematological laboratory data from many patients limits our ability to comment on whether the mechanism of CVT after COVID-19 is likely to be similar or different from that observed after ChAdOx1 nCoV-19 or Ad26.COV2.S. In particular, we do not have information regarding anti-platelet factor 4 (PF4) antibodies that have been associated with VITT [2,3].

In summary, COVID-19 is associated with a markedly increased incidence of CVT compared to patients with influenza, people who have received BNT162b2 or mRNA-1273 vaccines and compared to the best estimates of the general population incidence. The risk with COVID-19 also appears greater than with ChAdOx1 nCoV-19 and Ad26.COV2.S vaccines, although as noted this conclusion is indirect and tentative. The rarity of CVT in all populations means that larger sample sizes are required to confirm the results, and complementary study designs are needed to aid interpretation. Nevertheless, the current data highlight the risk of serious thrombotic events in COVID-19, and can help contextualize and inform debate about the risk-benefit ratio for current COVID-19 vaccines.

## Data Availability

The TriNetX system returned the results of the analyses as .csv files, which were downloaded and archived. Data presented will be freely accessible at: [the URL will be added upon publication]. In addition, TriNetX will grant access to researchers if they have a specific concern (through a third-party agreement option).

## Acknowledgments

PJH and MT were granted unrestricted access to TriNetX Analytics for the purposes of research, and with no restrictions as to the analyses done or the decision to publish. We thank Prakash Bhuyan, Dennis Briley, Sue Pavord, Andy Pollard and Mary Ramsay for advice and helpful comments.

## Transparency declaration

The corresponding author is the guarantor and affirms that this manuscript is an honest, accurate, and transparent account of the study being reported; that no important aspects of the study have been omitted; and that any discrepancies from the study as planned have been explained.

## Copyright/License for publication

The Corresponding Author has the right to grant on behalf of all authors and does grant on behalf of all authors, a worldwide licence to the Publishers and its licensees in perpetuity, in all forms, formats and media (whether known now or created in the future), to i) publish, reproduce, distribute, display and store the Contribution, ii) translate the Contribution into other languages, create adaptations, reprints, include within collections and create summaries, extracts and/or, abstracts of the Contribution, iii) create any other derivative work(s) based on the Contribution, iv) to exploit all subsidiary rights in the Contribution, v) the inclusion of electronic links from the Contribution to third party material where-ever it may be located; and, vi) licence any third party to do any or all of the above.

## Dissemination declaration

We plan to disseminate the results to the public and patients and family members of patients with COVID-19.

