## Supplement for "Cerebral venous thrombosis and portal vein thrombosis: a retrospective cohort study of 537,913 COVID-19 cases"

### Supplementary methods

#### TriNetX network

This section provides a version of our previous description of the network. [1]

*Legal and ethical status*
TriNetX's Analytics network is compliant with the Health Insurance Portability and Accountability Act (HIPAA), the US federal law which protects the privacy and security of healthcare data. TriNetX is certified to the ISO 27001:2013 standard and maintains an Information Security Management System (ISMS) to ensure the protection of the healthcare data it has access to and to meet the requirements of the HIPAA Security Rule. Any data displayed on the TriNetX Platform in aggregate form, or any patient level data provided in a data set generated by the TriNetX Platform, only contains de-identified data as per the de-identification standard defined in Section §164.514(a) of the HIPAA Privacy Rule. The process by which the data is de-identified is attested to through a formal determination by a qualified expert as defined in Section §164.514(b)(1) of the HIPAA Privacy Rule. This formal determination by a qualified expert, refreshed in December 2020, supersedes the need for TriNetX's previous waiver from the Western Institutional Review Board (IRB). The network contains data that are provided by participating Health Care Organizations (HCOs), each of which represents and warrants that it has all necessary rights, consents, approvals and authority to provide the data to TriNetX under a Business Associate Agreement (BAA), so long as their name remains anonymous as a data source and their data are utilized for research purposes. The data shared through the TriNetX Platform are attenuated to ensure that they do not include sufficient information to facilitate the determination of which HCO contributed which specific information about a patient.

*Acquisition of data, quality control, and other procedures*
The data are stored onboard a TriNetX appliance – a physical server residing at the institution's data centre or a virtual hosted appliance. The TriNetX platform is a fleet of these appliances connected into a federated network able to broadcast queries to each appliance. Results are subsequently collected and aggregated.

Once the data are sent to the network, they are mapped to a standard and controlled set of clinical terminologies and undergo a data quality assessment including 'data cleaning' that rejects records which do not meet the TriNetX quality standards. HIPAA compliance of the clinical patient data is achieved using de-identification. Different data modalities are available in the network. They include demographics (coded to HL7 version 3 administrative standards), diagnoses (represented by ICD-10-CM codes), procedures (coded in ICD-10-PCS or CPT), and measurements (coded to LOINC). While extensive information is provided about patients' diagnoses and procedures, other variables (such as socioeconomic and lifetime factors) are not comprehensively represented.

The data from a typical HCO generally go back around 7 years, with some going back 13 years. The data are continuously updated. HCOs update their data at various times, with most refreshing every 1, 2, or 4 weeks.

The data come primarily (>93%) from HCOs in the USA, with the remainder coming from India, Australia, Malaysia, Taiwan, Spain, UK, and Bulgaria. Only 1.8% of patients with COVID-19 are contributed from HCOs outside the USA. As noted above, to comply with legal frameworks and ethical guidelines guarding against data re-identification, the identity of participating HCOs and their individual contribution to each dataset are not disclosed to researchers.

Data quality assessment followed a standardised strategy wherein the data are reviewed for conformance (adherence to specified standards and formats), completeness (quantifying data presence or absence) and plausibility (believability of the data from a clinical perspective). There are pre-defined metrics for each of the above assessment categories. Results for these metrics are visualised and reviewed for each new site that joins the network as well as on an ongoing basis. Any identified issue is communicated to the data provider and resolved before continuing data collection.

The basic formatting of contributed data is also checked (e.g. to ensure that dates are properly represented). Records are checked against a list of required fields (e.g., patient identifier) and rejects those records for which the required information is missing. Referential integrity checking is done to ensure that data spanning multiple database tables can be successfully joined together. As the data are refreshed, changes in volume of data over time is monitored to ensure data validity. At least one non-demographic fact for each patient is required for them to be counted in the dataset. Patient records with only demographics information are discarded.

The software also undergoes quality control. The engineers testing the software are independent from the engineers developing it. Each test code is checked by two independent testing engineers. Each piece of software is tested extensively against a range of synthetic data (i.e. generated for the purpose of testing) for which the expected output is established independently. If the software fails to return this output, then the software is deemed to have failed the test and is examined and modified accordingly. For statistical software (including that used for propensity score matching, for Kaplan-Meier analysis, etc), an additional quality control step is implemented. Two independent codes are written in two different programming languages (typically R and python) and the statistical results are compared. If discrepancies are identified, then the codes are deemed to have failed the test and are examined and modified accordingly. All the code is reviewed independently by another engineer.

The test strategy follows three levels of granularity:

1. Unit tests: These test specific blocks, or units, of code that perform specific actions (e.g. querying the database).
2. Integration tests: These ensure that different components are working together correctly.
3. End-to-end tests: These tests run the entire system and check the final output.

*Some comments on advantages and disadvantages of EHR data*

The advantage of EHR data, like those in TriNetX, over insurance claim data is that both insured and uninsured patients are included. An advantage of EHR data over survey data is that they represent the diagnostic rates in the population presenting to healthcare facilities. This provides an accurate account of the burden of specific diagnoses on healthcare systems. The downside of relying on diagnoses is that they obviously do not account for undiagnosed patients who might be suffering from the illness but did not seek medical attention (or in whom the diagnosis was missed). A general limitation of EHR data is that a patient may be seen in different HCOs for different parts of their care and if one HCO is not part of the federated network then part of their medical records may not be available. Using a network of HCOs (rather than a single HCO) limits this possibility but does not fully remove it. Finally, historical data before the start of EHRs (or the addition of an HCO to the network) may be incomplete.

**Cohorts definition and index events**

The two control cohorts used consisted of patients who received an mRNA vaccine and patients with a diagnosis of influenza. Specifically, patients who received the vaccine were those who had any of the following procedure codes in their electronic health records:

- 91300: "Severe acute respiratory syndrome coronavirus 2 (SARS-CoV-2) (Coronavirus disease [COVID-19]) vaccine, mRNA-LNP, spike protein, preservative free, 30 mcg/0.3mL dosage, diluent reconstituted, for intramuscular use"
- 0001A: "Immunization administration by intramuscular injection of severe acute respiratory syndrome coronavirus 2 (SARS-CoV-2) (Coronavirus disease [COVID-19]) vaccine, mRNA-LNP, spike protein, preservative free, 30 mcg/0.3mL dosage, diluent reconstituted; first dose"
- 0002A: "Immunization administration by intramuscular injection of severe acute respiratory syndrome coronavirus 2 (SARS-CoV-2) (Coronavirus disease [COVID-19]) vaccine, mRNA-LNP, spike protein, preservative free, 30 mcg/0.3mL dosage, diluent reconstituted; second dose"
- 91301: "Severe acute respiratory syndrome coronavirus 2 (SARS-CoV-2) (Coronavirus disease [COVID-19]) vaccine, mRNA-LNP, spike protein, preservative free, 100 mcg/0.5mL dosage, for intramuscular use"
- 0011A: "Immunization administration by intramuscular injection of severe acute respiratory syndrome coronavirus 2 (SARS-CoV-2) (Coronavirus disease [COVID-19]) vaccine, mRNA-LNP, spike protein, preservative free, 100 mcg/0.5mL dosage; first dose"
- 0012A: "Immunization administration by intramuscular injection of severe acute respiratory syndrome coronavirus 2 (SARS-CoV-2) (Coronavirus disease [COVID-19]) vaccine, mRNA-LNP, spike protein, preservative free, 100 mcg/0.5mL
- 2468231: "SARS-CoV-2 (COVID-19) vaccine, mRNA spike protein"

Patients with influenza were those who had any of the following diagnoses:

- J09: Influenza due to certain identified influenza viruses
- J10: Influenza due to other identified influenza virus
- J11: Influenza due to unidentified influenza virus.

Because some patients with the control index event might have had COVID-19 at a different point in time, we excluded from the control cohorts all those who had COVID-19 at any point in time. To avoid any contamination between cohorts, COVID-19 as an exclusion criterion was defined in the broader sense to be all patients with a confirmed diagnosis of COVID-19 (ICD-10 code U07.1) but also patients with an unconfirmed COVID-19 diagnosis (U07.2), a recorded positive PCR test for COVID-19, or any of the following recorded on or after January 20, 2020: Pneumonia due to SARS-associated coronavirus (J12.81), Other coronavirus as the cause of disease classified elsewhere (B97.29), or Coronavirus infection unspecified (B34.2). Inclusion of the latter three diagnostic codes captures patients who receive a COVID-19 diagnosis in the early stage of the pandemic when the ICD code for COVID-19 (U07) was not yet defined. Specifically, the following codes were excluded from the control cohorts if they occurred on or after January 20, 2020:

- U07.1: COVID-19, virus identified
- U07.2: COVID-19, virus not identified
- J12.81: Pneumonia due to SARS-associated coronavirus
- B97.29: Other coronavirus as the cause of disease classified elsewhere
- B34.2: Coronavirus infection, unspecified
- Positive SARS-CoV-2 RNA in Respiratory specimen
- Positive SARS-CoV-2 RNA in Unspecified specimen
- Positive SARS-CoV-2 N gene in Respiratory specimen
- Positive SARS-CoV-2 N gene in Unspecified specimen
- Positive SARS-CoV-2 RdRp gene in Respiratory specimen
- Positive SARS-CoV-2 E gene in Respiratory specimen
- Positive SARS-CoV-2 E gene in Unspecified specimen
- Positive SARS-CoV-2 RNA panel in Respiratory specimen
- Positive SARS-CoV-2 RNA panel in Unspecified specimen
- Positive SARS-CoV-2 RNA in Nasopharynx
- Positive SARS coronavirus 2 and related RNA
- Positive SARS-related coronavirus RNA in Respiratory specimen
- Positive SARS coronavirus 2 ORF1ab in Respiratory specimen

**Baseline characteristics code**

When reporting baseline characteristics, the following ICD-10 codes are used:

- Obesity: E66
- Hypertension: I10-I16
- Chronic kidney disease: N18
- Ischemic heart disease: I20-I25
- Heart failure: I50
- Disease of the arteries, arterioles, or capillaries: I70-I79
- Disease of (non-cerebral) veins: I80-I87
- Cerebral/Pre-cerebral artery stenosis/occlusion: I63 (cerebral infarction), I65 (Occlusion and stenosis of precerebral arteries, not resulting in cerebral infarction), I66 (Occlusion and stenosis of cerebral arteries, not resulting in cerebral infarction)
- Intracranial hemorrhage: I60 (Nontraumatic subarachnoid hemorrhage), I61 (Nontraumatic intracerebral hemorrhage), I62 (Other and unspecified nontraumatic intracranial hemorrhage)
- Dementia: F01 (Vascular dementia), F02 (Dementia in other diseases classified elsewhere), F03 (Unspecified dementia), G30 (Alzheimer's disease), G31.0 (Frontotemporal dementia), and G31.83 (Dementia with Lewy bodies)
- Chronic lower respiratory diseases: J40-J47

- Connective tissue disorders: M30-M36
- Liver diseases: K70-K77
- Diabetes mellitus: E08-E13
- Malignancy: C00-C14 (Malignant neoplasms of lip, oral cavity and pharynx), C15-C26 (Malignant neoplasms of digestive organs), C30-C39 (Malignant neoplasms of respiratory and intrathoracic organs), C40-C41 (Malignant neoplasms of bone and articular cartilage), C43-C44 (Melanoma and other malignant neoplasms of skin), C45-C49 (Malignant neoplasms of mesothelial and soft tissue), C50 (Malignant neoplasms of breast), C51-C58 (Malignant neoplasms of female genital organs), C60-C63 (Malignant neoplasms of male genital organs), C64-C68 (Malignant neoplasms of urinary tract), C69-C72 (Malignant neoplasms of eye, brain and other parts of central nervous system), C73-C75 (Malignant neoplasms of thyroid and other endocrine glands), C76-C80 (Malignant neoplasms of ill-defined, other secondary and unspecified sites), C7A (Malignant neuroendocrine tumors), C7B (Secondary neuroendocrine tumors), C81-C96 (Malignant neoplasms of lymphoid, hematopoietic and related tissue)

**Details of statistical analysis**

In propensity score matching, the propensity score was calculated using a logistic regression (implemented by the function LogisticRegression of the scikit-learn package in Python 3.7). To eliminate the influence of ordering of records, the order of the records in the covariate matrix were randomised before matching.

### Supplementary figures

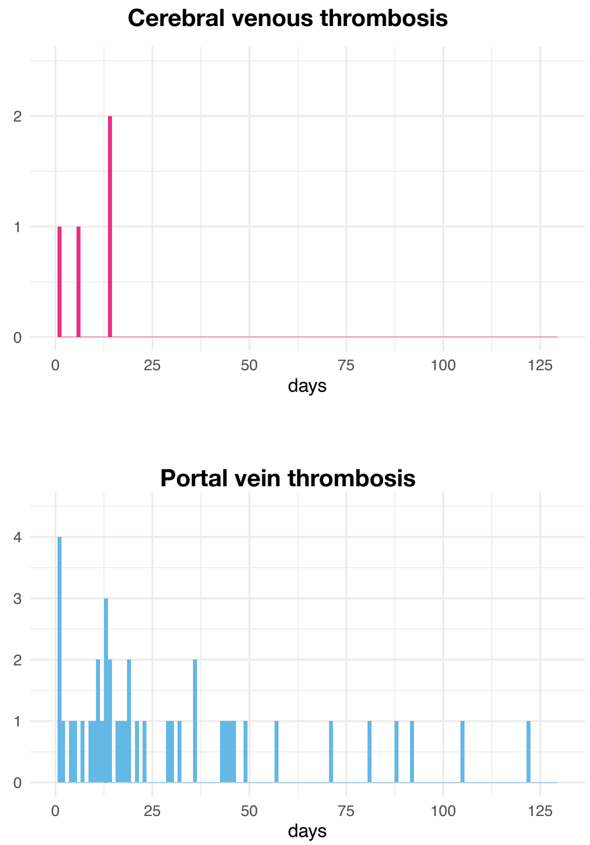


**Fig. S1** – Distribution of the day of recorded death relative to the diagnosis of CVT/PVT for patients who died after having had a CVT post COVID-19 (top) or a PVT post COVID-19 (bottom).

### Supplementary table

**Table S1** – Baseline characteristics of the matched cohorts of patients diagnosed with COVID-19 vs. influenza. SMD=Standardized mean difference.

| | COVID-19 | Influenza | SMD |
|---|---|---|---|
| **Sample size**, n | 392424 | 392424 | - |
| **Age**, mean (SD), y | 40.9 (20.7) | 41.3 (21.0) | 0.02 |
| **Sex**, n (%) | | | |
| Female | 218345 (55.6) | 231499 (59.0) | 0.07 |
| Male | 173970 (44.3) | 160872 (41.0) | 0.07 |
| Other | 109 (0.03) | 53 (0.01) | 0.01 |
| **Race,** n (%) | | | |
| White | 242092 (61.7) | 258548 (65.9) | 0.09 |
| Black or African American | 67373 (17.2) | 65270 (16.6) | 0.01 |
| Asian | 11110 (2.8) | 11573 (2.9) | 0.007 |
| American Indian or Alaska Native | 1653 (0.4) | 1622 (0.4) | 0.001 |
| Native Hawaiian or Other Pacific Islander | 1083 (0.3) | 1291 (0.3) | 0.01 |
| Unknown | 69113 (17.6) | 54120 (13.8) | 0.1 |

**Table S2** – Baseline characteristics of the matched cohorts of patients diagnosed with COVID-19 and people receiving an mRNA vaccine. SMD=Standardized mean difference.

| | COVID-19 | mRNA Vaccine | SMD |
|---|---|---|---|
| **Sample size**, n | 366869 | 366869 | - |
| **Age**, mean (SD), y | 55.2 (18.1) | 55.0 (18.1) | 0.01 |
| **Sex**, n (%) | | | |
| Female | 207968 (56.7) | 210226 (57.3) | 0.01 |
| Male | 158734 (43.3) | 156536 (42.7) | 0.01 |
| Other | 167 (0.05) | 107 (0.03) | 0.008 |
| **Race,** n (%) | | | |
| White | 248653 (67.8) | 245477 (66.9) | 0.02 |
| Black or African American | 52633 (14.3) | 52069 (14.2) | 0.004 |
| Asian | 13385 (3.6) | 16959 (4.6) | 0.05 |
| American Indian or Alaska Native | 1878 (0.5) | 2042 (0.6) | 0.006 |
| Native Hawaiian or Other Pacific Islander | 718 (0.2) | 746 (0.2) | 0.002 |
| Unknown | 49602 (13.5) | 49576 (13.5) | 2.00E-04 |

STROBE Statement—Checklist of items that should be included in reports of ***cohort studies***

| | Item No | Recommendation |
|---|---|---|
| **Title and abstract** | 1 | (*a*) Indicate the study's design with a commonly used term in the title or the abstract |
| | | (*b*) Provide in the abstract an informative and balanced summary of what was done and what was found |
| **Introduction** | | |
| Background/rationale | 2 | Explain the scientific background and rationale for the investigation being reported |
| Objectives | 3 | State specific objectives, including any prespecified hypotheses |
| **Methods** | | |
| Study design | 4 | Present key elements of study design early in the paper |
| Setting | 5 | Describe the setting, locations, and relevant dates, including periods of recruitment, exposure, follow-up, and data collection |
| Participants | 6 | (*a*) Give the eligibility criteria, and the sources and methods of selection of participants. Describe methods of follow-up |
| | | (*b*) For matched studies, give matching criteria and number of exposed and unexposed |
| Variables | 7 | Clearly define all outcomes, exposures, predictors, potential confounders, and effect modifiers. Give diagnostic criteria, if applicable |
| Data sources/ measurement | 8* | For each variable of interest, give sources of data and details of methods of assessment (measurement). Describe comparability of assessment methods if there is more than one group |
| Bias | 9 | Describe any efforts to address potential sources of bias |
| Study size | 10 | Explain how the study size was arrived at |
| Quantitative variables | 11 | Explain how quantitative variables were handled in the analyses. If applicable, describe which groupings were chosen and why |
| Statistical methods | 12 | (*a*) Describe all statistical methods, including those used to control for confounding |
| | | (*b*) Describe any methods used to examine subgroups and interactions |
| | | (*c*) Explain how missing data were addressed |
| | | (*d*) If applicable, explain how loss to follow-up was addressed |
| | | (*e*) Describe any sensitivity analyses |
| **Results** | | |
| Participants | 13* | (a) Report numbers of individuals at each stage of study—eg numbers potentially eligible, examined for eligibility, confirmed eligible, included in the study, completing follow-up, and analysed |
| | | (b) Give reasons for non-participation at each stage |
| | | (c) Consider use of a flow diagram |
| Descriptive data | 14* | (a) Give characteristics of study participants (eg demographic, clinical, social) and information on exposures and potential confounders |
| | | (b) Indicate number of participants with missing data for each variable of interest |
| | | (c) Summarise follow-up time (eg, average and total amount) |
| Outcome data | 15* | Report numbers of outcome events or summary measures over time |
| Main results | 16 | (*a*) Give unadjusted estimates and, if applicable, confounder-adjusted estimates and their precision (eg, 95% confidence interval). Make clear which confounders were adjusted for and why they were included |
| | | (*b*) Report category boundaries when continuous variables were categorized |
| | | (*c*) If relevant, consider translating estimates of relative risk into absolute risk for a meaningful time period |

| | | |
|---|---|---|
| Other analyses | 17 | Report other analyses done—eg analyses of subgroups and interactions, and sensitivity analyses |
| **Discussion** | | |
| Key results | 18 | Summarise key results with reference to study objectives |
| Limitations | 19 | Discuss limitations of the study, taking into account sources of potential bias or imprecision. Discuss both direction and magnitude of any potential bias |
| Interpretation | 20 | Give a cautious overall interpretation of results considering objectives, limitations, multiplicity of analyses, results from similar studies, and other relevant evidence |
| Generalisability | 21 | Discuss the generalisability (external validity) of the study results |
| **Other information** | | |
| Funding | 22 | Give the source of funding and the role of the funders for the present study and, if applicable, for the original study on which the present article is based |

*Give information separately for exposed and unexposed groups.

**Note:** An Explanation and Elaboration article discusses each checklist item and gives methodological background and published examples of transparent reporting. The STROBE checklist is best used in conjunction with this article (freely available on the Web sites of PLoS Medicine at http://www.plosmedicine.org/, Annals of Internal Medicine at http://www.annals.org/, and Epidemiology at http://www.epidem.com/). Information on the STROBE Initiative is available at http://www.strobe-statement.org.
